# Risk of Ischemic Stroke after COVID-19 Bivalent Booster Vaccination in an Integrated Health System

**DOI:** 10.1101/2023.09.11.23295368

**Authors:** Katie A Sharff, Thomas K Tandy, Paul F Lewis, Eric S Johnson

## Abstract

**Purpose:** What is the absolute occurrence of ischemic stroke and transient ischemic attack after a COVID-19 bivalent vaccination?

**Methods:** We conducted a retrospective cohort study of Kaiser Permanente Northwest (KPNW) patients 18 years and older who were vaccinated with either the Pfizer or Moderna formulation of the COVID-19 bivalent vaccine between September 1, 2022 and March 1^st^ 2023. Patients were included in the study if they had KP membership at the time of vaccination and through the 21-day follow up period. We replicated the Vaccine Safety Datalink (VSD) rapid cycle analysis methodology and searched for possible cases of ischemic stroke or TIA in the 21 days following vaccination using ICD-10-CM diagnosis codes in both the primary position and any position. We waited 90 days from the end of the follow up (March 21, 2023) for complete non-KP data accrual before analyzing the data to account for the lag in processing outside hospital insurance claims. Two physicians adjudicated possible cases by reviewing the clinical notes in the electronic health record. The analyses were stratified by age ≥65 years to allow for comparisons with VSD’s reporting at the Advisory Committee on Immunization Practices (ACIP) meeting of incidence of ischemic stroke or TIA (VSD reported incidence; 24.6 cases of ischemic stroke or TIA per 100,000 patients vaccinated).

**Results:** The incidence of ischemic stroke or TIA was 34.3 per 100,000 (95% CI, 17.7 – 59.9) in patients 65 years or older who received the bivalent Pfizer vaccine—based on a diagnosis code in the primary position of the emergency department or hospital discharge. The incidence increased to 45.7 per 100,000 (95% CI 26.1 – 74.2) when we expanded the search to a diagnosis in any position and did not adjudicate to confirm. However, most of those additional apparent stroke or TIA diagnoses were false-positive diagnoses based on physicians’ adjudications. Estimating the incidence based on the primary position agreed closely with estimating the incidence based on any position and physician adjudication: 37.1 per 100,000 (95% CI 19.8 – 63.5). Seventy-nine percent of the ischemic stroke cases were admitted to hospitals that are not owned by the integrated delivery system.

**Conclusion:** We identified a 50% increase in the incidence of ischemic stroke per 100,000 patients ages 65 and older vaccinated with the Pfizer bivalent vaccine, compared to the data presented by the VSD. Seventy-nine percent of the ischemic stroke cases were admitted to non-plan hospitals and a delay in processing outside hospital insurance claims was likely responsible for the discrepancy in case ascertainment of ischemic stroke. Physician adjudication of all cases in this study allowed accurate absolute incidence estimates of stroke per 100,000 vaccine recipients and is helpful in calculation of net benefit for policy recommendations and shared decision-making.

**5 key points:**

1. We identified a 50% increase in incidence of ischemic stroke or transient ischemic attack (TIA) per 100,000 patients ages 65 and older vaccinated with the Pfizer bivalent vaccine, compared to the data presented by the Vaccine Safety Datalink (VSD).
2. Seventy-nine percent of the ischemic stroke and TIA cases were admitted to hospitals that are not owned by the integrated delivery system.
3. A delay in processing outside hospital insurance claims was likely responsible for the discrepancy in case ascertainment of ischemic stroke.
4. Replication of the VSD case definition confirmed the exceptionally high positive predictive value (PPV) in identifying ischemic stroke or TIA within 21 days of Pfizer bivalent vaccination in individuals 65 years and older when the ICD-10-CM hospital discharge code is restricted to the primary position.
5. Physician adjudication of all cases in this study allowed accurate absolute incidence estimates of stroke per 100,000 vaccine recipients and is helpful in calculation of net benefit for policy recommendations and shared decision-making.

## Purpose

Bivalent Omicron BA4/5 COVID19 boosters were approved for use in the United States on August 31, 2022.^1^ The Centers for Disease Control (CDC) tracks safety data related to vaccines through the Vaccine Safety Datalink (VSD) which is a collaborative project between CDC and thirteen integrated healthcare systems in the US, including Kaiser Permanente Northwest (KPNW). The VSD utilizes rapid cycle analysis (RCA) methodology for near real-time surveillance of 21 prespecified outcomes during weekly sequential monitoring after COVID-19 bivalent booster vaccination.^2^

VSD’s RCA found a statistical signal (excess rate ratio) for ischemic stroke and TIA in individuals 65 years and older who received the bivalent Pfizer-BioNTech (Pfizer) COVID19 vaccine. Although the VSD identified an initial signal, the small number of strokes and imprecise rate ratios limited some analyses, and this signal was not replicated in other safety systems. ^3,4,5^

To estimate the incidence of ischemic stroke and TIA after a COVID-19 bivalent vaccination, we conducted a retrospective cohort study at KPNW. Because all potential ICD-10-CM-coded ischemic strokes would be adjudicated by physicians to exclude false-positive diagnoses, we searched for ICD-10-CM diagnosis codes in any position on a hospital discharge to improve sensitivity (complete case ascertainment). Our cohort study addresses a gap in the evidence: We estimated the absolute occurrence of ischemic stroke to support improved diagnosis and calculation of net benefit for policy recommendations and shared decision-making.

## Methods

We conducted a retrospective cohort study of KPNW patients 18 years and older who were vaccinated with either the Pfizer or Moderna formulation of the COVID-19 bivalent vaccine between September 1, 2022 and March 1 2023. Patients were included in the study if they had KP membership at the time of vaccination and through the 21-day follow up period. We replicated the VSD RCA methodology and searched for possible cases of ischemic stroke or TIA in the 21 days following vaccination. The following ICD-10-CM diagnosis codes were included to find the incident cases: G45.8, G45.9 and I63*. The primary diagnosis was defined by an incident case ICD-10-CM code in the primary diagnosis field of the hospital discharge: emergency department visits or hospital admissions. The VSD’s RCA protocol and publications do not specify whether they restricted their search to the primary position for ICD-10-CM diagnosis codes for emergency department visits or hospitalizations, but the primary position seems likely given the high PPV and lack of routine adjudication of events. Furthermore, we waited 90 days from the end of the follow up (March 21, 2023) for complete non-KP data accrual before analyzing the data to account for the lag in processing outside hospital insurance claims. Patients were excluded if they had atrial fibrillation, sickle cell anemia, primary thrombophilia, prior history of TIA, or sequelae of cerebrovascular disease at any point prior to the incident case, COVID-19 within the past 30 days, acute MI within the past 28 days, injury of blood vessels at neck level, or arterial embolism/thrombosis within one day of the incident case as outlined in the VSD protocol.^2^ Two physicians adjudicated possible cases by reviewing the clinical notes in the electronic health record. The analyses were stratified by age ≥65 years to allow for comparisons with VSD’s reporting at the Advisory Committee on Immunization Practices (ACIP) meeting (24.6 cases of ischemic stroke or TIA per 100,000 patients vaccinated).^4^ For the incidence estimates and positive predictive value estimates we calculated exact 95% confidence intervals using Stata 17 and the default Clopper-Pearson binomial method.^6^ The Institutional Review Board at KPNW approved this study.

## Results

We followed 105,171 patients 18 years and older vaccinated with at least one dose of Pfizer or Moderna COVID19 bivalent vaccines: 85,535 (81.3%) received Pfizer and 19,688 (18.7%) received by Moderna. When we restricted our search to the primary position of ICD-10-CM diagnosis code we identified 19 patients who met the VSD criteria for acute ischemic stroke or TIA within 21 days of vaccination. Two physicians confirmed that 18 of the 19 possible cases met the clinical definition of TIA or ischemic stroke, positive predictive value of 94.7% (95% CI, 74.0-99.9).

When we expanded our search to include an ICD-10-CM diagnosis code in any position (i.e., not restricted to the primary position) we identified 24 possible cases. Two physicians confirmed that 19 of the 24 cases met the clinical definition of TIA or ischemic stroke (Table). The expanded search with ICD-10-CM diagnosis codes in any position had a PPV of 79.2 % (95% CI, 57.8-92.9). The 5 cases that did not meet clinical adjudication had a variety of alternative diagnosis including dizziness or sepsis.

**Table:** Adjudicated ischemic stroke or TIA cases in all patients 18 years and older with ICD-10-CM hospital discharge diagnosis code in any position.

| Case | Age | Claims | Sex | Diagnosis | Vaccine Brand | Days from Vaccination to Onset of Stroke/TIA | High Dose Flu Vaccine | Timing of High Dose Flu Vaccine |
| --- | --- | --- | --- | --- | --- | --- | --- | --- |
| 1 | 40-49 | Yes | Male | R middle cerebral artery occlusion | Pfizer | 12 | No | NA |
| 2 | 50-59 | Yes | Male | L middle cerebral artery ischemic stroke | Pfizer | 20 | No | NA |
| 3 | 50-59 | Yes | Female | Lacunar Infarct | Pfizer | 11 | No | NA |
| 4 | 50-59 | Yes | Male | Subacute Infarct R frontal lobe | Pfizer | 21 | No | NA |
| 5 | 60-69 | Yes | Female | TIA | Pfizer | 7 | No | NA |
| 6 | 60-69 | Yes | Male | TIA | Pfizer | 14 | No | NA |
| 7 | 60-69 | Yes | Male | Infarct of R ventral pons | Pfizer | 5 | Yes | Same day |
| 8 | 60-69 | Yes | Male | R thalamic infarct | Pfizer | 21 | Yes | Same Day |
| 9 | 70-79 | Yes | Female | Acute Ischemic stroke | Moderna | 10 | Yes | Same day |
| 10 | 70-79 | Yes | Female | Acute Pontine Infarct | Pfizer | 21 | Yes | Same Day |
| 11 | 70-79 | Yes | Male | Stroke in pons | Pfizer | 6 | Yes | 21 days prior |
| 12 | 70-79 | Yes | Male | Expressive aphasia, presumptive stroke, unable to image | Pfizer | 4 | Yes | >30 days prior |
| 13 | 70-79 | Yes | Male | TIA | Pfizer | 15 | Yes | >30 days prior |
| 14 | 70-79 | Yes | Female | Acute multifocal L hemisphere stroke | Pfizer | 11 | Yes | >30 days prior |
| 15 | 80-89 | Yes | Male | TIA followed by infarct post-central gyrus | Pfizer | 5 | Yes | 20 days prior |
| 16 | 80-89 | Yes | Female | Acute microvascular infarcts | Pfizer | 0 | No | NA |
| 17 | 80-89 | Yes | Male | TIA | Pfizer | 18 | Yes | Same day |
| 18 | 80-89 | Yes | Female | Right basal ganglia stroke | Pfizer | 10 | Yes | >30 days prior |
| 19 | 80-89 | Yes | Male | R basal ganglia ischemic stroke | Pfizer | 1 | Yes | Same Day |

*TIA: Transient ischemic attack*
*R: Right*
*L: Left*
*NA: Not applicable*
*Pfizer: Pfizer-BioNTech COVID-19 Vaccine, Bivalent*
*Moderna: Moderna COVID-19 Vaccine Bivalent*

When we restricted the analysis to patients 65 years and older who received the bivalent Pfizer vaccine—based on the VSD’s RCA signal detection--we identified 12 cases with an ICD-10-CM in the primary position and all were confirmed: PPV=100% (95% CI, 73.5-100). When we expanded the search to any position, there were 16 cases and 13 were confirmed by adjudication: PPV=81.3% (95% CI, 54.4-96.0).

When we restricted the analysis to the 35,003 patients 65 years and older who received the bivalent Pfizer vaccine, we identified an incidence of ischemic stroke or TIA of 34.3 per 100,000 (95% CI, 17.7 – 59.9) based on the primary code position. For the same group of patients the incidence increased when a diagnosis could appear in any position: 45.7 per 100,000 (95% CI, 26.1-74.2)(non-adjudicated); 37.1 per 100,000 (95% CI, 19.8-63.5) (adjudicated and confirmed).

## Conclusion

The incidence of ischemic stroke or TIA was 34.3 per 100,000 (95% CI, 17.7 – 59.9) in patients 65 years or older who received the bivalent Pfizer vaccine—based on a diagnosis code in the primary position of the emergency department or hospital discharge. The incidence increased to 45.7 per 100,000 (95% CI, 26.1 – 74.2) when we expanded the search to a diagnosis in any position and did not adjudicate to confirm. However, most of those additional apparent stroke or TIA diagnoses were false-positive diagnoses based on physicians’ adjudications. Estimating the incidence based on the primary position agreed closely with estimating the incidence based on any position and physician adjudication: 37.1 per 100,000 (95% CI, 19.8 – 63.5), which is a 50% increase in incidence as presented by the VSD at ACIP for this same population (24.6 cases per 100,000 patients vaccinated (non-adjudicated). ^4^

Our findings demonstrated that the current VSD protocol has a high PPV 94.7 (95% CI, 74.0-99.9) in identifying ischemic stroke or TIA within 21 days of bivalent booster vaccine in patients ages 18 and older, and a PPV=100% (95% CI, 73.5-100) in Pfizer recipients ages 65 and older. Our findings corroborate the VSD estimate presented at the ACIP meeting in which they reported a PPV of 92% for Pfizer vaccine recipients ages 65 and older.^3^

Bivalent COVID-19 vaccine first became available in the United States on August 31, 2023.^7^ The VSD identified a safety signal of ischemic stroke with clustering in the first 11-22 days after bivalent BNT162b2 Pfizer-BioNTech vaccination with a relative risk of 1.47 (95% CI 1.11-1.95) in the days 1-21 post vaccination compared to days 22-44 post-vaccination, however this signal was not replicated in other systems. ^3,4,5^

A possible explanation for our higher incidence relative to the VSD data may be related to the delay in processing outside hospitals’ insurance claims.^8^ In our analysis, 79% of the adjudicated cases were admitted to an outside hospital and we waited a full 90 days after the follow up period (March 21, 2023) to allow for complete case ascertainment. When insurance claims are used to identify inpatient diagnoses, claims delays are lengthy for sites that do not own the hospitals used by their patients.^9^ The VSD relies on real-time data with analysis updated weekly, but within the VSD system, at least 4 of the health systems rely on claims from hospitals that are not owned by the integrated delivery system.^3,4^

The position of the ICD-10-CM hospital discharge code (primary position vs any position) identified one additional case after physician adjudication and therefore was a small contribution to the discrepancy for case ascertainment of ischemic stroke between our study and the VSD’s rapid cycle analysis.

A limitation to this study is we were unable to account for underlying differences in the KPNW VSD population (age, gender, comorbid conditions) in comparison to the aggregate VSD population that may increase their inherent risk of stroke or TIA.

Because the rapid cycle surveillance requires weekly analyses of ICD-10-CM-coded emergency department visits and hospital admissions, that timeline allow insufficient time for physician adjudication through chart review; the case definitions for the 21 outcomes are designed to optimize the positive predictive value, sometimes at the expense of sensitivity (i.e., complete outcome ascertainment). The VSD’s rapid cycle case definitions support accurate rate ratios that inform timely signal detection. The VSD’s rapid cycle case definitions do not necessarily support accurate absolute incidence estimates that answer a distinct question: How often does the adverse event occur in patients who were recently vaccinated? Physicians need to know how often an adverse event occurs to reduce missed diagnoses—”Bayesian thinking.”^10^ Knowing how often an adverse event occurs also informs the calculation of net benefit for policy recommendations and shared decision-making.^11^

Our study identified a 50% increase in the incidence of ischemic stroke or TIA per 100,000 Pfizer bivalent vaccine recipients ages 65 and older compared to the data shared by VSD. The reasons for this finding are uncertain but may be related to a delay in processing of outside hospital claims.

## Data Availability

All data produced in the present study are available upon reasonable request to the authors

